# Biallelic variants in the non-coding RNA gene *RNU4-2* cause a recessive neurodevelopmental syndrome with distinct white matter changes

**DOI:** 10.1101/2025.08.13.25333306

**Authors:** Rocio Rius, Alexander JM Blakes, Yuyang Chen, Joachim De Jonghe, Javeria R Alvi, Florence Amblard, Christina Austin-Tse, Sarah Baer, Meena Balasubramanian, Elsa V Balton, Giulia Barcia, Jonathan A Bernstein, Pierre Blanc, Daniel Buchzik, Daniel G Calame, Benjamin Cogne, Charles Coutton, Chloe A Cunningham, Nitsuh Dargie, Christel Depienne, Katrina M Dipple, Anne Dieux, Abhijit Dixit, Lauren Dreyer, Haowei Du, Salima El Chehadeh, Michael Field, Vanessa Geiger, Richard A Gibbs, Ian Glass, Olivier Grunewald, Paul Gueguen, Tobias B Haack, Hamza Hadj Abdallah, Radu Harbuz, Bertrand Isidor, Marie-Line Jacquemont, Médéric Jeanne, G. Christoph Korenke, Urania Kotzaeridou, Richard J Leventer, James R Lupski, Pierre Marijon, Kaitlin E McGinnis, Rodrigo Mendez, Olfa Messaoud, Caroline Nava, Mevyn Nizard, Anne O’Donnell-Luria, Melanie C O’Leary, Simone Olivieri, Amitav Parida, Davut Pehlivan, Jennifer E Posey, Chloe M Reuter, Véronique Satre, Caroline Schluth-Bolard, Thomas Smol, Tipu Sultan, Christel Thauvin, Julien Thevenon, Eloise Uebergang, Catherine Vincent-Delorme, Evangeline Wassmer, Matthew T Wheeler, Elif Yilmaz Gulec, Adeline Vanderver, Arastoo Vossough, Stephan J Sanders, Siddharth Banka, Gregory M Findlay, Daniel G MacArthur, Cas Simons, Nicola Whiffin

## Abstract

Genetic variants in *RNU4-2*, which encodes U4, a key non-coding small nuclear RNA (snRNA) component of the major spliceosome, were recently shown to cause a prevalent neurodevelopmental disorder (NDD) called ReNU syndrome. These variants, which almost exclusively arise *de novo*, act in a dominant fashion and are clustered within 18 nucleotides (nt) in the centre of *RNU4-2*. Here we describe a novel recessive NDD associated with homozygous and compound heterozygous variants in *RNU4-2*. We identified 32 individuals with biallelic variants outside of the 18 nt ReNU syndrome region, that cluster within other functionally important elements of U4, including the Stem II region, the k-turn motif, and the Sm protein binding site. We characterise the clinical phenotype in 27 of these individuals, demonstrating that the recessive disorder is clinically distinct from dominant ReNU syndrome and is associated with distinctive white matter abnormalities, including enlarged perivascular spaces. Together, these findings expand the genotypic and phenotypic spectrum of *RNU4-2*-associated NDDs.

## INTRODUCTION

Splicing is a core cellular process mediated by a large ribonucleoprotein (RNP) complex called the spliceosome, which is composed of small nuclear RNAs (snRNAs) and numerous associated proteins^1^. The major spliceosome is responsible for catalysing the removal of ∼99.5% of introns through U2-dependent splicing, while the minor spliceosome is responsible for the remaining 0.5% which are U12-type introns^2^. Genetic variants in many components of both the major and minor spliceosomes cause a heterogenous group of disorders that are collectively termed “spliceosomopathies”^3^.

While the majority of spliceosomopathies discovered to date are caused by variants in the protein components of the spliceosome, an increasing number involve snRNAs. The first to be identified were recessive disorders caused by variants in the minor spliceosome snRNAs *RNU4ATAC*^4^, and *RNU12*^5,6^. More recently, *de novo* variants in *RNU4-2*, which encodes the U4 snRNA of the major spliceosome, were shown to cause ReNU syndrome, a dominant syndromic neurodevelopmental disorder (NDD)^7,8^. Subsequent to the discovery of ReNU syndrome, variants in two additional major spliceosomal snRNAs, *RNU2-2* and *RNU5B-1* were also identified to cause dominant NDDs^9–11^. Further, variants in *RNU4-2* and U6 encoding genes were shown to cause dominant isolated retinitis pigmentosa^12^. Across all of the spliceosomopathies, ReNU syndrome is the most prevalent, predicted to correspond to ∼0.4% of all severe NDD, or around 100,000 individuals worldwide^7^.

In a companion paper^13^, we describe a saturation genome editing (SGE) experiment, where we simultaneously measured the functional impact of variants across *RNU4-2* in a haploid cell line. These data led us to identify a novel recessive NDD caused by biallelic variants outside of the T-loop and Stem III regions in which heterozygous variants cause ReNU syndrome. These biallelic variants map to regions of U4 that are important for binding to other spliceosome components: the Stem II region of interaction with U6, the Sm protein binding site which is essential for snRNA biogenesis and stability, and the k-turn structure in the 5’ stem loop which binds to SNU13/15.5k. Strikingly, variants in the equivalent regions and nucleotides of *RNU4ATAC* (the minor spliceosome homologue of *RNU4-2*) cause the RNU4atac-opathies microcephalic osteodysplastic primordial dwarfism type I/III (MOPDI or Taybi-Linder syndrome; MIM 210710), Lowry–Wood syndrome (MIM 226960), and Roifman syndrome (MIM 616651)^13,14^.

Here, we describe the clinical phenotype of this novel autosomal recessive NDD caused by variants in *RNU4-2*. We identify a cohort of 32 individuals with biallelic variants in specific regions of *RNU4-2* and characterise the clinical phenotype of the recessive NDD syndrome in a subset of 27 individuals. We show that while this recessive disorder has phenotypic similarities with dominant ReNU syndrome, some features, including specific white matter changes, are distinct. These findings expand the phenotypic and genotypic spectrum of NDDs associated with *RNU4-2*, with *RNU4-2* being the first snRNA to be linked to both dominant and recessive disorders. Further, we establish recessive disease and variant features critical for facilitating accurate diagnosis.

## RESULTS

### Biallelic variants in RNU4-2 cause a syndromic neurodevelopmental disorder with distinct white matter abnormalities

Our companion manuscript describes an SGE experiment where we measured the functional impact of variants across *RNU4-2*^13^. Each variant (n=539) was given a ‘function score’ as a measure of the depletion of cells with the variant across two time points. Variants with a function score < −0.39 were determined to be significantly depleted and hence to have an effect on cell viability through impacting *RNU4-2* function. Initially, we searched rare disease cohorts for undiagnosed individuals with biallelic variants with significant SGE function scores in regions of *RNU4-2* not yet associated with NDD^13^. This resulted in identification of 19 individuals from 13 families; 10 individuals (including 3 pairs of siblings) with homozygous variants and 9 individuals (including 3 pairs of siblings) with compound heterozygous variants. We also identified one individual in Genomics England who was classified as diagnosed, but who was compound heterozygous for two *RNU4-2* variants with significant SGE function scores (individual 30; see below).

In this study, we expanded our analysis to search for additional undiagnosed NDD families with biallelic variants in *RNU4-2* in global rare disease cohorts (see **Methods**), including those with non-significant SGE scores. In total, we identified 38 individuals across 30 families (**Sup. Table 1**). Thirteen individuals from ten families had homozygous variants and 25 individuals from 20 families had compound heterozygous variants. Nine individuals from six families, all of whom had homozygous variants, were reported to have consanguineous parents. In the Genomics England 100,000 genomes project, we identified biallelic *RNU4-2* variants in 5/5,386 trios with undiagnosed NDD, compared to 0/4,776 trios with non-NDD phenotypes (one-sided Fisher’s *P*=0.042).

**Table 1:** Summary of phenotypes observed in 27 individuals with biallelic *RNU4-2* variants. Only phenotypes observed in at least 20% of individuals are included. Separate counts are shown for (1) individuals with biallelic P/LPvariants, (2) individuals where one or both alleles had VUS, and (3) all individuals.

|  | **Both Alleles P/LP**<br>**% (Present/Known)** | **Other**<br>**% (Present/Known)** | **Total**<br>**% (Present/Known)** |
| --- | --- | --- | --- |
| **Age last seen (Median, range)** | 10y, 4-32 | 10y, 2-21 | 10y, 2-32 |
| **Sex** | 9M, 2F | 10M, 6F | 19M, 8F |
| **Global developmental delay (GDD)** | 100.0% (11/11) | 100.0% (16/16) | 100.0% (27/27) |
| GDD severe | 63.6% (7/11) | 46.7% (7/15) | 53.8% (14/26) |
| GDD moderate | 36.4% (4/11) | 53.3% (8/15) | 46.2% (12/26) |
| **Intellectual disability (ID)** | 100.0% (11/11) | 100.0% (16/16) | 100.0% (27/27) |
| ID Severe | 54.5% (6/11) | 46.7% (7/15) | 50.0% (13/26) |
| ID Moderate | 36.4% (4/11) | 46.7% (7/15) | 42.3% (11/26) |
| ID Mild | 9.1% (1/11) | 6.7% (1/15) | 7.7% (2/26) |
| **Impaired language development** | 100.0% (11/11) | 100.0% (16/16) | 100.0% (27/27) |
| Absent speech | 63.6% (7/11) | 50.0% (8/16) | 55.6% (15/27) |
| **Gross motor delay / delayed walking** | 100.0% (11/11) | 93.8% (15/16) | 96.3% (26/27) |
| Inability to walk >5y | 30.0% (3/10) | 30.8% (4/13) | 30.4% (7/23) |
| **Abnormality of brain MRI** | 100.0% (10/10) | 78.6% (11/14) | 87.5% (21/24) |
| Abnormality of cerebral white matter | 100.0% (10/10) | 78.6% (11/14) | 87.5% (21/24) |
| Dilated perivascular spaces | 100.0% (8/8) | 75.0% (9/12) | 85.0% (17/20) |
| Abnormality of the corpus callosum | 37.5% (3/8) | 40.0% (4/10) | 38.9% (7/18) |
| Abnormality of the cerebellum | 40.0% (4/10) | 30.0% (3/10) | 35.0% (7/20) |
| Hypotonia | 81.8% (9/11) | 87.5% (14/16) | 85.2% (23/27) |
| Dysmorphic facial features | 54.5% (6/11) | 93.8% (15/16) | 77.8% (21/27) |
| **Abnormality of the eye** | 72.7% (8/11) | 81.2% (13/16) | 77.8% (21/27) |
| Strabismus/ Esotropia | 33.3% (3/9) | 62.5% (10/16) | 52.0% (13/25) |
| Seizure | 72.7% (8/11) | 56.2% (9/16) | 63.0% (17/27) |
| Behavioral abnormality | 42.9% (3/7) | 64.3% (9/14) | 57.1% (12/21) |
| Spasticity | 45.5% (5/11) | 56.2% (9/16) | 51.9% (14/27) |
| Abnormal integument | 45.5% (5/11) | 53.3% (8/15) | 50.0% (13/26) |
| Microcephaly | 40.0% (4/10) | 50.0% (8/16) | 46.2% (12/26) |
| Feeding difficulties | 55.6% (5/9) | 38.5% (5/13) | 45.5% (10/22) |
| Short stature | 10.0% (1/10) | 62.5% (10/16) | 42.3% (11/26) |
| **Abnormality of Movement / Coordination** | 54.5% (6/11) | 31.2% (5/16) | 40.7% (11/27) |
| Dental anomalies | 22.2% (2/9) | 50.0% (7/14) | 39.1% (9/23) |
| Abnormality of the genitalia | 36.4% (4/11) | 26.7% (4/15) | 30.8% (8/26) |
| Abnormality of the skeletal system | 27.3% (3/11) | 33.3% (5/15) | 30.8% (8/26) |
| Endocrine anomalies | 27.3% (3/11) | 33.3% (5/15) | 30.8% (8/26) |
| ASD | 11.1% (1/9) | 44.4% (4/9) | 27.8% (5/18) |
| Failure to thrive | 11.1% (1/9) | 35.7% (5/14) | 26.1% (6/23) |

We identified only eleven individuals in the UK Biobank with biallelic variants in *RNU4-2*. Five of the 490,541 genome sequenced participants had homozygous variants and six individuals from a subset of 200,011 participants with phased genome sequencing data^15^ had compound heterozygous variants (**Sup. Table. 2**). Of the 14 unique variants observed in the UK Biobank individuals in the homozygous or compound heterozygous state, only one has a significant SGE score (n.120T>C, SGE=-1.15).

For each individual in the UK Biobank and NDD cohort, we calculated the mean SGE function score for the variants identified on their two alleles. Individuals with biallelic variants in the UK Biobank had significantly weaker mean SGE scores than individuals with NDD (UKBiobank mean= −0.101; NDD mean= −0.498; two-sided Mann Whitney U test *P*=1.3×10^−4^; **Fig. 1**). We excluded NDD individuals with SGE scores similar to those observed in the general population from further characterisation, using a threshold of −0.2 that maximally separated individuals in the UK Biobank from those with NDD (**Fig. 1**). This threshold provides a distinction for analysis but should be interpreted as a pragmatic case definition rather than definitive (see **Discussion**). This led to a cohort of 32 individuals from 24 families that we used for all following analyses (**Sup. Table 1**).

**Figure 1:**
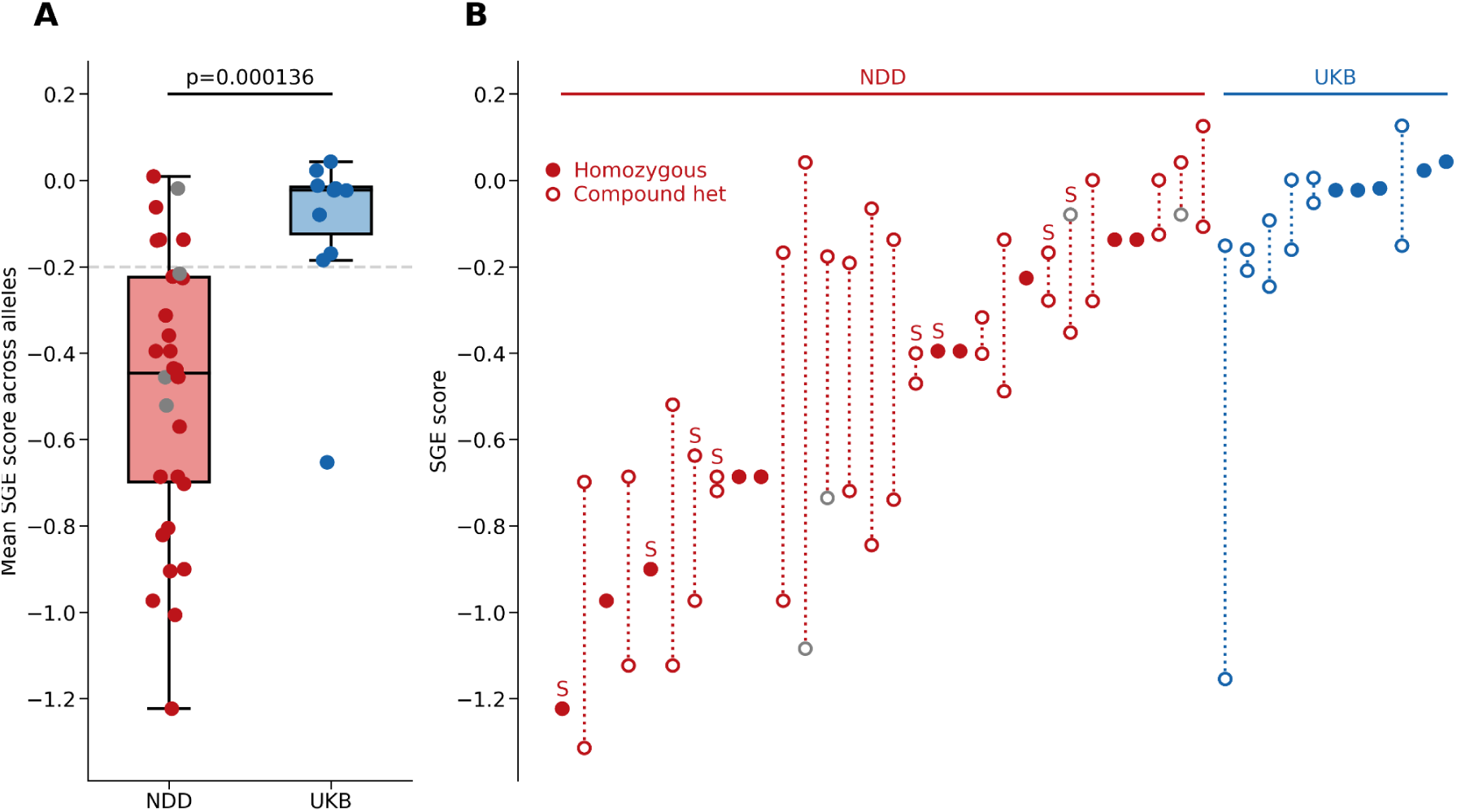
SGE function scores for individuals with biallelic *RNU4-2* variant with NDD (red) versus individuals in UK Biobank (blue). (**A**) Mean SGE scores across both alleles per individual. Only one individual from each sibling pair was included. Box and whisker plots show the median, quartiles, and +/− 1.5 times the interquartile range of the data. Mean scores were compared with a two-sided Mann Whitney U test. The SGE score cutoff (mean SGE score −0.2) used to include participants in the characterised NDD cohort is shown with a grey dashed line. (**B**) SGE scores per allele for each individual. Variants from sibling pairs are annotated with the letter “S”). Across both A and B, for insertions and deletions without available SGE data the SGE score was inferred from the mean SGE score across all SNVs within the deleted nucleotides or taking the mean SGE score across all SNVs within the nucleotides directly flanking the insertion. Individuals (panel A) and variants (panel B) with these inferred scores are shown in grey.

### Clinical characterisation of the biallelic RNU4-2 associated NDD

Of the 32 included individuals with biallelic *RNU4-2* variants, detailed clinical data were available for 27 (19 males, 8 females) through contact with their clinical teams (**Fig. 2A**; **Table 1**; **Sup. Table 4**). The median age at last follow-up was 10 years (range: 2–32 years). Most individuals had infantile onset phenotypes (n=15; 55.5%), while seven (25.9%) had congenital/neonatal onset and five (18.5%) had childhood onset.

**Fig. 2:**
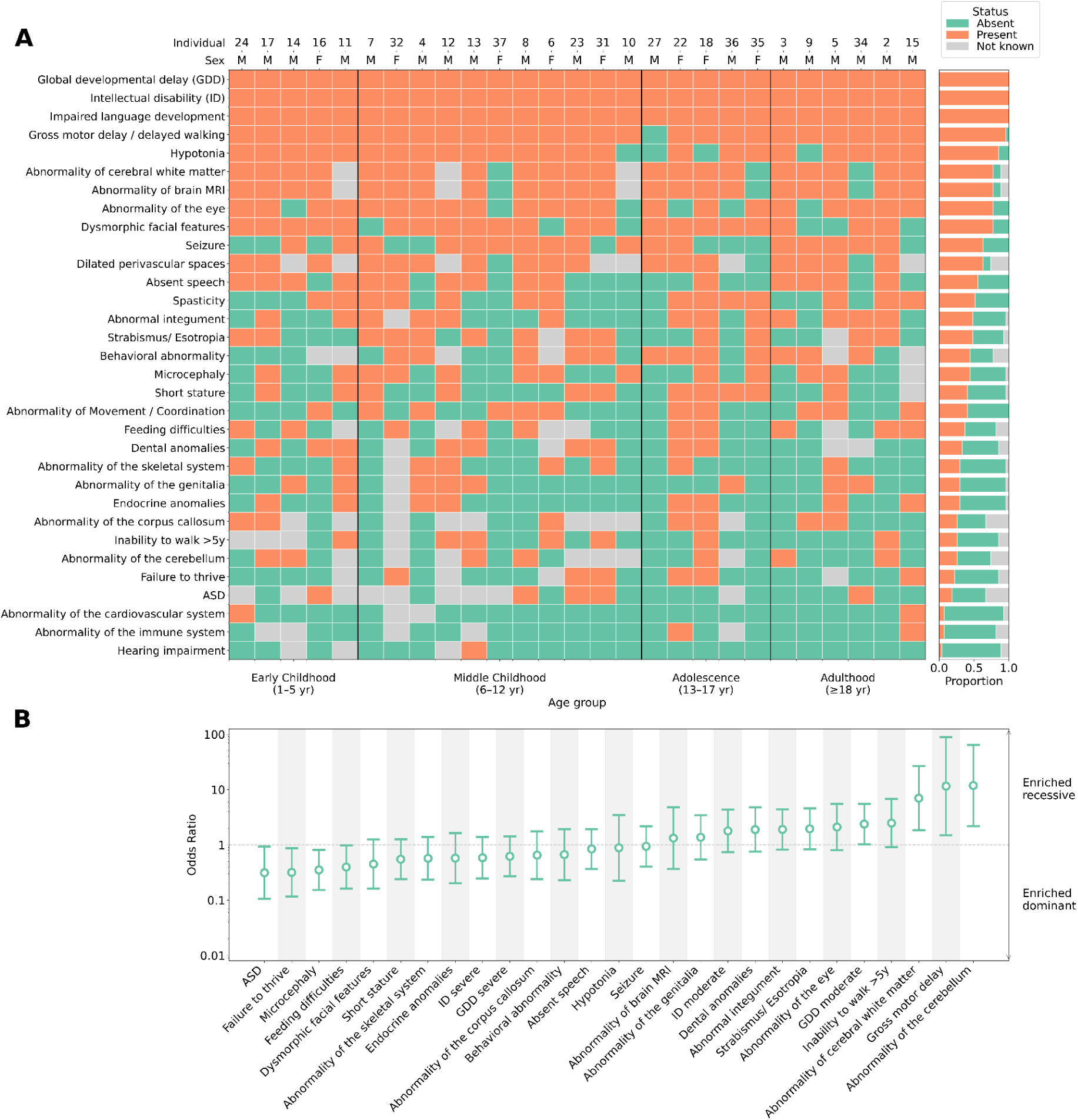
Clinical features of individuals with biallelic *RNU4-2* variants. **(A)** Colour map illustrating Human Phenotype Ontology (HPO) terms present in 27 individuals with the recessive *RNU4-2* NDD and detailed clinical data available. Individuals are stratified by age at last evaluation. The sex and individual number (used throughout the text and in Sup. Tables 1 and 4) are shown across the top. (**B**) Phenotype enrichment or depletion in 27 individuals with the recessive *RNU4-2* NDD versus 178 individuals with dominant ReNU syndrome. Phenotypes are limited to those observed in at least 25% of individuals in this study or the combined ReNU cohorts. No phenotypes differed significantly between the cohorts after false discovery rate (FDR) correction (two-sided Z tests on log-transformed values). Error bars show 95% confidence intervals.

Global developmental delay (GDD) or intellectual disability (ID) was present in all individuals (n=27). Severity data, available for 26 individuals, revealed severe GDD in fourteen (53.8%), and moderate GDD in twelve (46.2%). All individuals (100%; 27/27) exhibited delayed language development. Fifteen (55.6%) individuals older than 2 years were non-verbal or had no meaningful spoken language. Of the twelve individuals with some expressive language, all reported delayed first words (median 3.8y, range: 20 months-8 years) and a limited expressive output, ranging from <10 words to simple sentences. Inability to walk was reported in 7/23 individuals above 5 years old (30.4%). Among the 17 individuals who achieved independent ambulation, first steps were consistently delayed, with a median of 2.5 years ranging from 1.5 years to over 5 years. Behavioural abnormalities were reported in 12/21 (57.1%), including obsessive-compulsive traits (n = 6), aggression or self-injurious behaviour (n = 6), and emotional lability, tantrums, or meltdowns (n = 6).

Most individuals had a history of hypotonia (85.2%; 23/27), with neonatal onset in 64% (16/25), which was commonly associated with feeding difficulties. Spasticity was present in 14/27 individuals (51.9%), while movement and coordination abnormalities were observed in 11/27 individuals (40.7%; six with ataxia, three with dystonia, one with choreoathetosis and one unspecified abnormality of coordination). Seizures occurred in 17/27 individuals (63%) with a median onset of 2.5 years. Seizure types at onset varied and included tonic-clonic, focal, generalized, febrile, absence, and startle-triggered seizures. In most cases, seizure semiology evolved over time. Of the 14 individuals with available data, 13 (92.8%) were treatment-responsive, while one had pharmacoresistant epilepsy with persistent daily seizures. No individuals were reported to have experienced status epilepticus.

Genital anomalies were observed in 8/19 males (42.1%), including micropenis, cryptorchidism, hypoplastic scrotum, and testicular ectopia. No genital abnormalities were reported in females (n=9). Heterogenous integumentary abnormalities were noted in 13 of 26 individuals (50.0%), with features such as hypertrichosis (n=4), livedo reticularis (n=2), acrokeratosis verruciformis (n=2), hypoplastic nails (n=2), hypopigmented macular lesions (n=2) and pigmentary changes (n=1).

Individual 30 was classified as diagnosed by Genomics England with a likely pathogenic variant in *GLI3* (NM_000168.6:c.804_810del, heterozygous). However, this *GLI3* variant did not explain all of their reported phenotypes, including white matter abnormalities and microcephaly. The absence of polydactyly in this individual was also inconsistent with *GLI3*-related disorder (MIM 175700 and 146510).

### Neuroimaging reveals consistent white matter involvement

Brain MRI data or reports were available for 24 individuals. Among these, 21 individuals showed abnormalities, most commonly including dilation of perivascular spaces in the periventricular white matter and white matter volume loss (**Sup. Table 4**). Only three individuals with imaging performed after one year of age were reported to have normal MRIs (Individuals 34, 35, and 37), although images were not available for review.

For 11 individuals, imaging data were directly reviewed by the same pediatric neuroradiologist, enabling detailed comparison (**Fig. 2, Sup. Table 4**). Cerebral white matter changes were the most consistent feature, present in 100% (11/11), most frequently manifesting as dilation of perivascular spaces in the periventricular and deep white matter. In more severe cases, this pattern resembled tightly packed microcysts (n=5; individuals 2, 3, 5, 9, and 32). Two individuals whose MRI was performed at <5 years old showed only minimal dilation (individuals 4 and 16). Two individuals (2 and 3) underwent serial imaging from infancy: initial scans showed only ventriculomegaly, but follow-up studies revealed progressive pathology, including perivascular space dilation, white matter volume loss, and cerebellar atrophy (**Fig. 2**; both ages <5 years). Individuals with the most severe dilation of perivascular spaces had lower mean SGE scores across their two variant alleles than individuals with minimal dilation evident on MRI review (−0.820 (SD 0.324) vs −0.390 (SD 0.192); *P*=0.0397; two-sided Mann Whitney U test with siblings excluded).

### Dominant and recessive RNU4-2 NDDs have distinct phenotypic features

We compared the phenotypes observed in 27 individuals with the recessive disorder to 178 individuals from two large non-overlapping studies of dominant ReNU syndrome (49 from Chen *et al*.^7^ and 129 from Nava *et al*.^11^). Many phenotypes are observed at similar frequencies across the recessive and dominant *RNU4-2* disorders, including global developmental delay (100% and 99.4%, respectively), marked speech and language delay (100% and 92.7%), and seizures (57.1% and 64.4%) (**Sup. Table 5**). Beyond neurodevelopmental phenotypes, the eye (77.8% and 62.5%) and skeletal system (30.8% and 43.8%) are commonly affected in both disorders.

However, the recessive disorder presents with distinct phenotypic features that allow clinical differentiation. Whereas non-specific white matter changes are common in ReNU syndrome (23/46; 50.0% in Chen *et al*.), the recessive disorder frequently presents dilated perivascular spaces that can mimic a compact microcystic appearance (17/20; 85.0%; **Fig. 2**) that have not been reported in ReNU syndrome. Cerebellar atrophy is observed in 35.0% (7/20) of the recessive individuals showing the largest raw enrichment in the recessive disorder, although this was not significant after multiple-testing correction (OR 11.85; 95%CI 2.19-64.13; unadjusted *P*=0.004; FDR-corrected *P*=0.060; two-sided Z test on log-transformed values; **Fig. 2B**). The prevalence of dysmorphic facial features was not significantly different between the two disorders (OR 0.45; 95%CI 0.16-1.25, FDR *P*=0.278), however, the reported dysmorphic features differ. Dominant ReNU syndrome usually features a myopathic facial appearance with deep-set eyes, epicanthal folds, a broad nasal bridge and anteverted nares, large cupped ears, full cheeks, a tented philtrum, and a triangular open mouth with full lips, downturned corners, and an everted lower lip vermilion^11^. In contrast, the dysmorphic features in recessive condition were variable, including high anterior hairline, synophrys, strabismus, upslanted palpebral features, broad nasal bridge and base, bulbous tip, a thin upper lip, and dental diastema (**Fig. 3**).

**Fig. 3:**
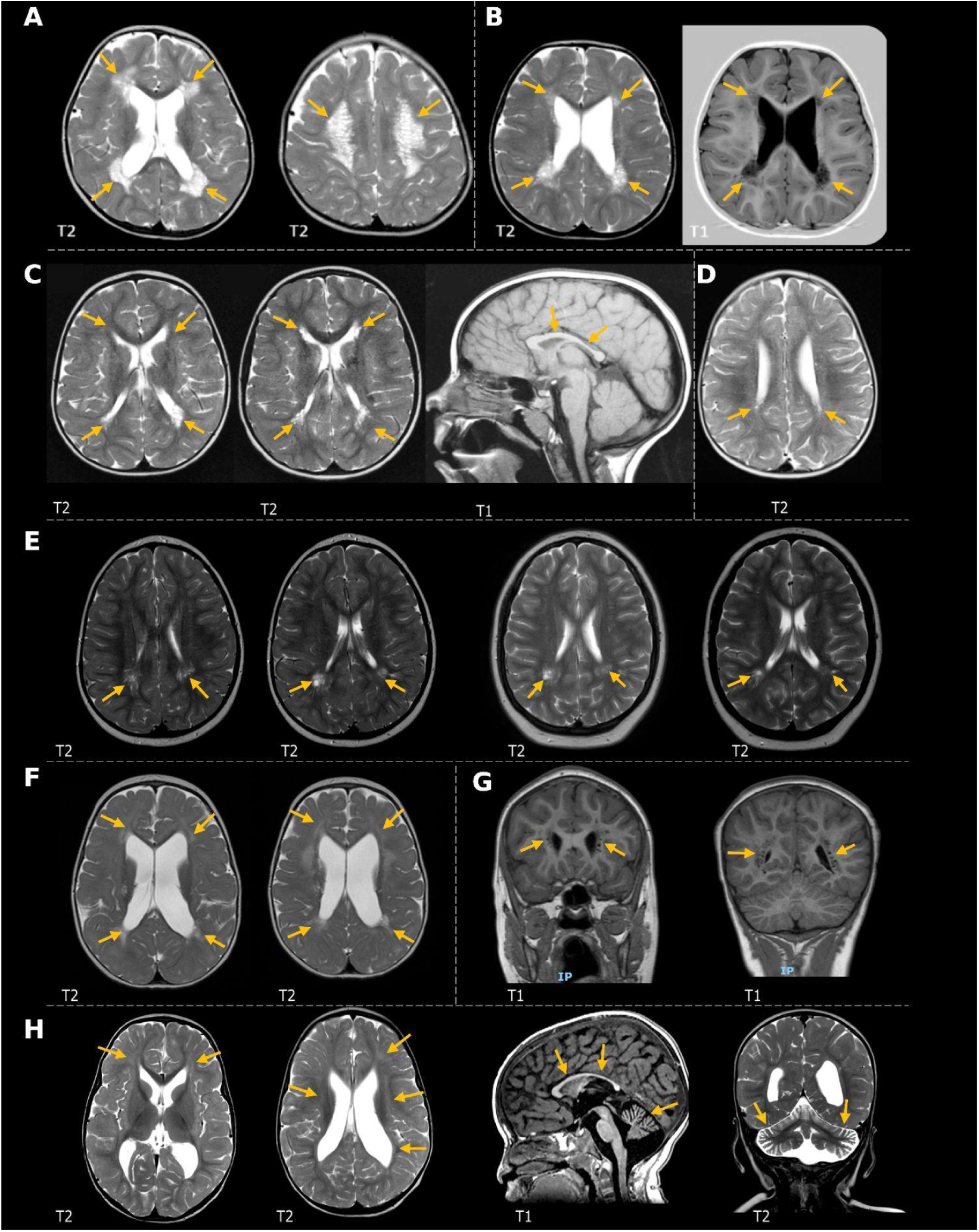
Individuals with biallelic *RNU4-2* variants have consistent cerebral white matter abnormalities. **(A, B)** Axial MRIs of individuals 2 and 3 show extensive dilated perivascular spaces in the periventricular region mimicking a tightly packed microcystic pattern and white matter volume loss. (**C**) In individual 22, the extent of dilated perivascular spaces slightly increased between scans at <5 years and 5-9 years, with T2-weighted axial views showing progression and a T1-weighted sagittal view at 5-9 years showing a thin corpus callosum. (**D**) T2-weighted axial view of individual 23, the sibling of individual 22, at <5 years shows less severe dilatation of the periventricular perivascular spaces. (**E**) T2-weighted axial views of individual 27 at ages <5 years and 10-14 years show periventricular focally dilated perivascular spaces in the peritrigonal region bilaterally. (**F**) T2-weighted axial view of individual 32 at <5 years shows periventricular dilated perivascular spaces, other patchy areas of white matter signal abnormality, low volume of the white matter, and ventriculomegaly. (**G**) T1-weighted coronal view of individual 5 shows periventricular dilated perivascular spaces at 10-14 years. (**H**) Individual 17’s MRI (axial and coronal T2-weighted and sagittal T1-weighted images) at <5 years shows faint T2 hyperintensities in the white matter, low white matter volume, and ventriculomegaly along with a thin corpus callosum and atrophy of the cerebellum.

### Biallelic variants in RNU4-2 cluster in important regions revealed by SGE

In the 32 included individuals with recessive *RNU4-2* NDD, we identified 30 unique *RNU4-2* variants, including six unique homozygous variants in 11 individuals (including three sibling pairs) and 26 unique variants as compound heterozygous (**Fig. 4**). Two variants (n.119A>G and n.7G>C) were observed in both homozygous and compound heterozygous genotypes. While only 17/30 (56.7%) variants had significant function scores in the SGE assay (5/6 (83.3%) homozygous variants and 11/26 (42.3%) compound heterozygous variants), the other variants were located in the same regions of the U4 structure: eight in the k-turn / 5’ stem loop (61.5%), three in Stem II (23.1%), and two in the terminal stem loop (15.4%; **Fig. 4**). For 12/30 (40.0%) of the variants, a variant at the equivalent nucleotide of *RNU4ATAC* was pathogenic/likely pathogenic in ClinVar and/or in Benoit-Pilven *et al*.^14^ (including 6/13 (46.2%) variants with non-significant or absent SGE scores). For ten further variants (four with non-significant or absent SGE scores), there was another variant identified in a different individual either at the same nucleotide (e.g. n.7G>A and n.7_8insA) or a pairing nucleotide in the 5’ stem loop of the U4 structure (e.g. n.28C>G and n.45G>C; **Sup. Table 3**). Furthermore, the location of the UK Biobank variants within the U4 structure differed from those identified in individuals with NDD, with seven UK Biobank variants (50%) observed in the 3’ stem loop of *RNU4-2* (n.85 to n.117), compared to 0/30 variants in NDD cases (*P*=0.001, two-sided Fisher’s exact test; **Fig. 4**).

**Fig. 4:** Facial photographs of individuals with biallelic variants identified in *RNU4-2*. **(A)** Individual 2. (**B**) Individual 3 (sibling of **A**). (**C**) Individual 4 (profile and frontal views). (**D**) Individual 11. (**E**) individual 34. (**F**) Individual 27. (**G**) Individual 32. (**H**) Individual 35. Dysmorphic features were variable across individuals, but commonly included a high anterior hairline (seen in **A**,**E**,**F**,**G**), synophrys (**A**,**C**,**D**,**E**,**H**), strabismus (**A**,**C**,**D**,**E**,**F**,**G**) upslanted palpebral fissures (**D**,**E**,**F**,**G**), broad nasal bridge and base (**D**,**E**,**F**), thin upper lip (**A**,**F**,**G**), and dental diastema (**A**,**D**,**F**,**G**).

**Fig. 5:**
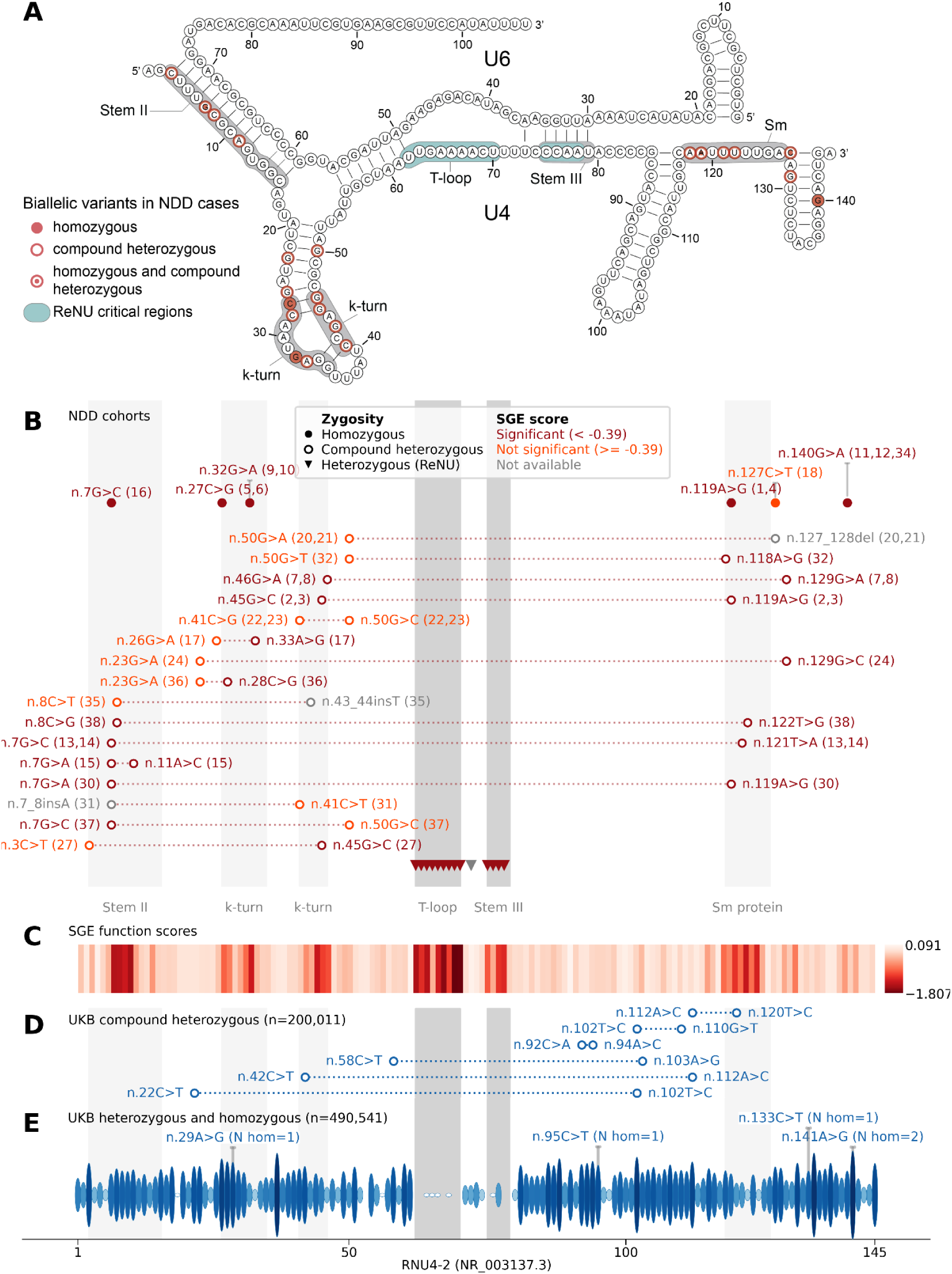
Biallelic variants identified in individuals with NDD cluster in structurally and functionally important regions of *RNU4-2*. **(A)** Schematic of U4 snRNA (*RNU4-2*, NR_003137.3) secondary structure in complex with U6. Variants are superimposed on affected nucleotides. (**B**) Variants identified in 37 individuals with NDD. Variants are coloured by their function score in the SGE assay^13^ with significantly depleted variants (function score < −0.39) in red, assayed but not significant variants (function score ≥ −0.39) in orange, and variants not included in the SGE assay in grey. The participant number (from **Sup. Tables 1 and 4**) associated with each variant, or combination of variants, is given in circular brackets. (**C**) Heatmap of SGE function scores. The minimum SGE score across all SNVs at each position is shown. (**D**) and (**E**) Variants observed in UK Biobank. (**D**) Compound heterozygous variants are shown from a subset of 200,011 individuals for whom statistical phasing data is available^15^. (**E**) Heterozygous and homozygous variants are shown for the full cohort of 490,541 genome sequenced individuals. For homozygous and heterozygous variants, the height of each ellipse is proportional to the logarithm of the allele count for the most frequent variant at that position (maximum allele count = 1,625). The number of homozygous individuals for each variant is shown. **Full plot:** Key structural regions are shaded in grey (the 5’ and 3’ stem loop regions and the Stem I region are not shown for clarity). The regions where ReNU syndrome variants occur are shown in teal (**A**) or darker grey (**B**-**E**). Regions important for snRNA-snRNA or snRNA-protein interactions where biallelic variants cluster are shaded in light grey. Labels for each region are shown above the SGE score heatmap (**C**) and in panel **A**. Short indels of 1-2nt are shown at their most 5’ position.

Using these regional and variant-level annotations and the ACMG/AMP framework, the 30 variants were curated (see **Methods**). Eleven variants (36.7%) reached a likely pathogenic classification, one pathogenic, and the rest remained a variant of uncertain significance (VUS). 14 of the 32 individuals (43.8%) had variants on both alleles that were classified as pathogenic or likely pathogenic, four had a single pathogenic or likely pathogenic allele, and the remaining 14 individuals had two alleles classified as VUS; **Sup. Table 3**). Across all clinical features, none occurred at a significantly different frequency in individuals with two pathogenic / likely pathogenic alleles when compared to all other individuals (**Table 1**).

For compound heterozygous individuals with the recessive disorder, we find no difference in mean SGE scores between maternally vs paternally inherited variants (−0.50 and −0.39 respectively, *P*=0.372, two-sided Mann Whitney U test). Similarly, in a compound heterozygote, the variant with the strongest SGE score is no more likely to be inherited maternally vs paternally (*P*=0.527, Chi squared goodness of fit test). We did not observe phenotypic clustering based on variant SGE scores, variant classification, or variant position within the U4 secondary structure (**Sup. Fig. 1**), however, these analyses are currently underpowered due to our limited sample size.

## DISCUSSION

Here, we characterise the clinical phenotype of a novel recessive NDD associated with variants in the *RNU4-2* snRNA gene. We show that this NDD is both genetically and phenotypically distinct from ReNU syndrome, which is caused by heterozygous variants within two critical regions in an 18 nt region in the centre *RNU4-2*. Pathogenic variants for the recessive *RNU4-2* condition fall outside the 18 nt ReNU region, instead clustering within other functionally important elements of *RNU4-2* including the Stem II region, k-turn region, and the Sm protein binding site.

The imaging findings in the recessive *RNU4-2*-associated disorder include cerebellar abnormalities and a prominent white matter phenotype on brain MRI. The most common pattern consists of dilated perivascular spaces, to varying degrees, in the periventricular and deep white matter regions, often accompanied by corpus callosum and cerebellar atrophy. Modern high-resolution MR imaging has improved detection of dilated perivascular spaces, which in mild forms can be benign^16^. In this cohort, several patients demonstrated very extensive and coalescent dilatation of perivascular spaces, producing a tightly packed microcystic appearance. While this severe MRI pattern appears characteristic of the recessive *RNU4-2* NDD, other individuals showed milder perivascular enlargement, overlapping with patterns reported in other NDDs^17–19^, and metabolic conditions^20^ with varying disease mechanisms.The presence of these changes on MRI, particularly the marked perivascular space dilation, should prompt sequencing and analysis of variants in *RNU4-2*.

ReNU syndrome is remarkably prevalent for an NDD, with a frequency similar to many well-known disorders caused by variants in large protein-coding genes^7^. In contrast, recessive *RNU4-2* associated NDD is much rarer. For example, while we identified 61 individuals with ReNU syndrome in 8,841 individuals (0.69%) with previously undiagnosed NDD in the Genomics England 100,000 Genomes Project, we only identified seven (0.08%) with biallelic variants in the same cohort. The relative frequencies of these disorders will, however, differ in populations with high rates of consanguinity, as evidenced by seven individuals (including 3 sibling pairs) with homozygous variants having consanguineous parents. *RNU4-2*, similar to other snRNA genes, has a substantially elevated mutation rate^21^. This, combined with negative selection acting on variants across the gene, results in a high density of variants across individuals, but all of these variants remain very rare^7^, lowering the chance of homozygous variants arising in populations with low levels of consanguinity.

We discovered this recessive *RNU4-2* associated NDD from an SGE experiment, which is detailed in a companion manuscript^13^. Using a stringent significance threshold, we initially identified 19 individuals from 13 families with biallelic variants that were significantly depleted in the SGE assay. In this study, we expand this cohort to include individuals with variants that do not meet the strict SGE significance threshold. Multiple lines of evidence support our assertion that the individuals with SGE significant and non-significant variants have the same recessive *RNU4-2* associated NDD: (1) a consistent phenotype, including observation of the same white matter anomalies on MRI, (2) the localisation of variants in each set of individuals to the same structural regions of the U4 snRNA, which differ from the localisation of variants in individuals in the UK Biobank, (3) variants with both significant and non-significant SGE scores occurring at the equivalent nucleotides as known pathogenic variants in *RNU4ATAC*, supporting that they are disruptive to U4 function.

While the SGE scores are highly predictive for dominant ReNU syndrome variants, they appear less sensitive for recessive disease. This may reflect differences in disease mechanisms: SGE scores measure cell fitness in a haploid cell line and may not capture all relevant aspects of *RNU4-2* function. As such, normal SGE scores should not be used as evidence of benignity in a recessive context.

Without a way to confidently distinguish deleterious from benign variation it is difficult to ascertain exactly which individuals should be characterised as having the recessive NDD. Here, we used the distribution of SGE scores of biallelic variants in the UK Biobank to determine a threshold for inclusion, defining a set of individuals with NDD falling outside this range for initial clinical characterisation. The consistent phenotype observed across many members of this cohort suggests that the majority of these individuals are in fact affected by recessive *RNU4-2* syndrome, but some may be false positives, and some of the excluded individuals may also have this recessive NDD. Larger characterised cohorts will be critical to better understand the spectrum of penetrance and variable expressivity for this syndrome and to define criteria to establish variant pathogenicity. Further, future assays using disease-relevant cellular models may aid the assessment of the functional impact of recessive variants in *RNU4-2*.

In summary, we have characterised the phenotype associated with biallelic variants in *RNU4-2* as a moderate to severe syndromic neurodevelopmental disorder with distinct white matter abnormalities. These data add to the phenotypic and genotypic spectrum of *RNU4-2* associated NDDs and highlight the increasing importance of screening snRNA genes to end the diagnostic odyssey for patients with undiagnosed NDD.

## METHODS

### NDD cohort and clinical data collection

We searched rare disease cohorts for individuals with biallelic variants in *RNU4-2* and undiagnosed neurodevelopmental phenotypes. These cohorts included the Genomics England (GEL) 100,000 Genomes Project and NHS Genomic Medicine Service datasets accessed through the UK National Genomic Research Library^22^, the Center for Population Genomics CaRDinal cohort, the SeqOIA and Auragen clinical cohorts in France (PFMG 2025; https://pfmg2025.fr/en/), the Undiagnosed Disease Network (UDN), the Broad Institute Center for Mendelian Genomics (CMG) and GREGoR (Genomics Research to Elucidate the Genetics of Rare Diseases)^23^ Consortium cohorts. Individuals were excluded if the variants didn’t segregate with NDD in the family. Variants were excluded if they were observed as homozygous in either UK Biobank or All of Us. Additional individuals were identified through personal communications. For individuals recruited as trios, variants were phased using parental sequencing data. For individuals with two variants in *RNU4-2* but without sequencing data from one or both parents, variant phasing was manually determined by inspection of reads in IGV^24^.

Informed consent was obtained for all patients included in this study from their parent(s) or legal guardian, with the study approved by the local regulatory authority. The 100,000 Genomes Project Protocol has ethical approval from the HRA Committee East of England Cambridge South (REC Ref 14/EE/1112).

Clinical collaboration requests were submitted to Genomics England to contact recruiting clinicians and collect additional phenotypic information. Clinical data were collected and summarised for features seen across the cohort. Written consent was obtained to publish all photographs and MRI images.

### Comparison of recessive phenotypes with dominant ReNU syndrome

For a subset of 27 individuals in this cohort for whom detailed phenotypic information was available (**Sup. Table 4**), we counted the number of individuals in whom each phenotype was present or definitely absent.

We obtained the same information for ReNU syndrome by combining counts from Table 1 and Supplementary Table 2 of Chen *et al*.^7^ and Supplementary Table 7 of Nava *et al*.^11^ given that the authors of Nava *et al*. took care to ensure that the two cohorts were non-overlapping. We included only phenotypes which were present in at least 25% of individuals with biallelic variants from this study, or at least 25% of individuals in the combined ReNU cohorts. Details of the precise phenotypic terms aggregated across the cohorts are given in **Sup. Table 5**. We then calculated the odds ratio for the presence of each phenotype in this cohort versus the combined ReNU cohorts. Statistical significance was determined by two-sided Z tests on the log-transformed values, followed by FDR correction (Benjamini-Hochberg method).

### Phenotypic clustering analysis

We reproduced the principal components analysis (PCA) described in Nava et al^11^. For individuals with detailed clinical phenotyping data, HPO terms were encoded as present (1) or absent (0). Missing data were annotated as absent. Intellectual disability was further stratified into mild (1), moderate (2), or severe (3) categories. PCA was performed on these encoded data. Only one individual from each sibling pair was included in this analysis.

### Annotation of biallelic variants in NDD cases and population controls

We identified variants in *RNU4-2* from short-read genome sequencing data in 490,541 individuals from the UK Biobank^25^ (DRAGEN pipeline) and in 414,840 individuals from All of Us V8. We additionally identified individuals with compound heterozygous variants in a subset of 200,011 UK Biobank participants with statistical phasing information^15^.

SGE scores were taken from De Jonghe *et al*.^13^. For Fig. 3, for each nucleotide position in *RNU4-2*, we calculated the minimum SGE score of any SNV at that position. To compare individuals with NDD to those in UK Biobank, for each individual with biallelic variants in *RNU4-2*, we calculated the mean SGE score across their two alleles.

Variants were annotated with the region of *RNU4-2* to which they map using the following nucleotides: Stem II (n.3 to n.16), 5’ stem loop (n.20 to n.52), Stem I (n.56 to n.62), t-loop (n.63 to n.70), Stem III (n.75 to n.79), 3’ stem loop (n.85 to n.117), Sm protein (n.118 to n.126), terminal stem loop (n.127 to n.144). Within the 5’ stem loop, the k-turn was annotated as n.27 to n.35 and n.41 to n.46.

Two regions of *RNU4-2* and *RNU4ATAC* with identical structures were defined as follows: *RNU4-2* n.26 to n.52 with *RNU4ATAC* n.31 to n.57 and *RNU4-2* n.115 to n.126 with *RNU4ATAC* n.113 to n.124). Variants at the same nucleotide in the structure and where the reference bases in *RNU4-2* and *RNU4ATAC* are identical, were marked as ‘equivalent’. Within the 5’ stem loop of *RNU4-2*, pairing nucleotides were determined as in Figure 4 of De Jonghe *et al*.^13^.

### Variant classification

All identified variants were classified using the established framework from the American College of Medical Genetics and Genomics (ACMG) and Association for Molecular Pathology (AMP)^26^ and additional specifications for non-coding variants^27^. ‘PM2 supporting’ was applied for variants rare in UK Biobank (AF<0.1% and no homozygotes). ‘PS3 supporting’ was applied to variants with significant SGE function scores, with the evidence level capped at supporting due to the absence of known pathogenic and benign variation to properly benchmark this assay for biallelic *RNU4-2* variants. ‘PM1’ was applied to variants in Stem II, the k-turn, or the Sm protein site (see above section). ‘PM3’ was applied following updated guidance from ClinGen (https://clinicalgenome.org/site/assets/files/3717/svi_proposal_for_pm3_criterion_-_version_ <u>1.pdf</u>) but with no evidence given to variants in trans with variants of uncertain significance given the high variant density across *RNU4-2*. ‘PM5’ was applied to variants with an exact equivalent variant in *RNU4ATAC* classified as Pathogenic or Likely Pathogenic in ClinVar. ‘PM5 supporting’ was applied for variants with a pathogenic/likely pathogenic variant at the same nucleotide within *RNU4-2*, or at a pairing nucleotide in a stem in the U4 structure.

## Data Availability

Research on the de-identified patient data used in this publication from the Genomics England 100,000 Genomes Project and the NHS GMS dataset can be carried out in the Genomics England Research Environment subject to a collaborative agreement that adheres to patient-led governance. All interested readers will be able to access the data in the same manner that the authors accessed the data. For more information about accessing the data, interested readers may contact or access the relevant information on the Genomics England website: https://www.genomicsengland.co.uk/research. Genomic and phenotypic data from the GREGoR consortium, including the RGP cohort, and the UDN are available through the dbGaP accession numbers phs003047.v1.p1 and phs001232.v5.p2, respectively, with at least annual data releases. Access is managed by a data access committee designated by dbGaP and is based on intended use of the requester and allowed use of the data submitter as defined by consent codes.

## ACKNOWLEDGEMENTS

We thank Peter O’Donovan, Mitra Sato and Eve Miller from the Genomics England Airlock team.

N.W. is supported by a Wellcome Career Development Award (grant no. 305292/Z/23/Z), a Lister Institute research prize, and grant funding from Novo Nordisk. Y.C. is supported by a studentship from Novo Nordisk. The Francis Crick Institute receives its core funding (G.M.F.) from Cancer Research UK (CC2190), the UK Medical Research Council (CC2190), and the Wellcome Trust (CC2190). A.B. is supported by a Wellcome PhD Training Fellowship for Clinicians and the 4Ward North PhD Programme for Health Professionals (223521/Z/21/Z). Analysis was supported by the Centre for Population Genomics (Garvan Institute of Medical Research and Murdoch Children’s Research Institute) and was funded in part by a National Health and Medical Research Council investigator grant (2009982) and the Medical Research Future Fund (MRFF) Genomics Health Futures Mission (2032931). Massimo’s Mission acknowledges funding support from the Australian Government Department of Health and Aged Care (EPCD000034). O.M. is supported by the Hazem Ben-Gacem Tunisia Medical Fellowship Fund. D.G.C. was supported by the National Institute of Neurological Disorders and Stroke of the National Institutes of Health under award number K12NS098482. This study was supported by the National Institute for Health and Care Research (NIHR) Manchester Biomedical Research Centre (NIHR203308) and funded in part by US National Institutes of Health Genomics Research Elucidates Genetics of Rare, GREGoR, Program (HG011758). Sequencing and analysis of Individual 24 were provided by the Broad Institute Center for Mendelian Genomics (Broad CMG) and were funded by the National Human Genome Research Institute (NHGRI) grants U01HG011755 (GREGoR consortium), R01HG009141, and in part by the Chan Zuckerberg Initiative Donor-Advised Fund at the Silicon Valley Community Foundation (funder DOI 10.13039/100014989) grants 2020-224274, 2022-309464, 2022-316726, and 2022-316726 (https://doi.org/10.37921/236582yuakxy). Research reported in this publication was supported by the National Institute Of Neurological Disorders And Stroke of the National Institutes of Health under Award Number U01NS134358. The content is solely the responsibility of the authors and does not necessarily represent the official views of the funding agencies.

This research was made possible through access to data in the National Genomic Research Library, which is managed by Genomics England Limited (a wholly owned company of the Department of Health and Social Care). The National Genomic Research Library holds data provided by patients and collected by the NHS as part of their care and data collected as part of their participation in research. The National Genomic Research Library is funded by the National Institute for Health Research and NHS England. The Wellcome Trust, Cancer Research UK and the Medical Research Council have also funded research infrastructure. This study was registered with Genomics England under Research Registry Projects 354. This research has been conducted using the UK Biobank Resource under application number 81050. We gratefully acknowledge All of Us and UK Biobank participants for their contributions. We also thank the National Institutes of Health’s All of Us Research Program for making available the participant and variant data examined in this study.

For the purpose of Open Access, the authors have applied a CC BY public copyright license to any Author Accepted Manuscript version arising from this submission.

## SUPPLEMENTARY FIGURES AND TABLE LEGENDS

**Supplementary Table 1:** Details of biallelic *RNU4-2* variants identified in individuals with NDD. AC = allele count.

**Supplementary Table 2:** Details of biallelic *RNU4-2* variants identified in individuals in the UK Biobank.

**Supplementary Table 3**: Details of individual variants identified in the 32 characterised individuals.

**Supplementary Table 4:** Detailed clinical information for 27 individuals with biallelic RNU4-2 variants. Y=Yes, N=No, NA=Not available/applicable.

Redacted for preprint; the table will be included in journal submission.

**Supplementary Table 5:** Phenotype enrichment analysis in 27 individuals with biallelic *RNU4-2* variants and 178 individuals with ReNU syndrome. Statistical significance was determined by two-sided Z tests on the log-transformed values, followed by FDR correction (Benjamini-Hochberg method).

**Supplementary Figure 1:**
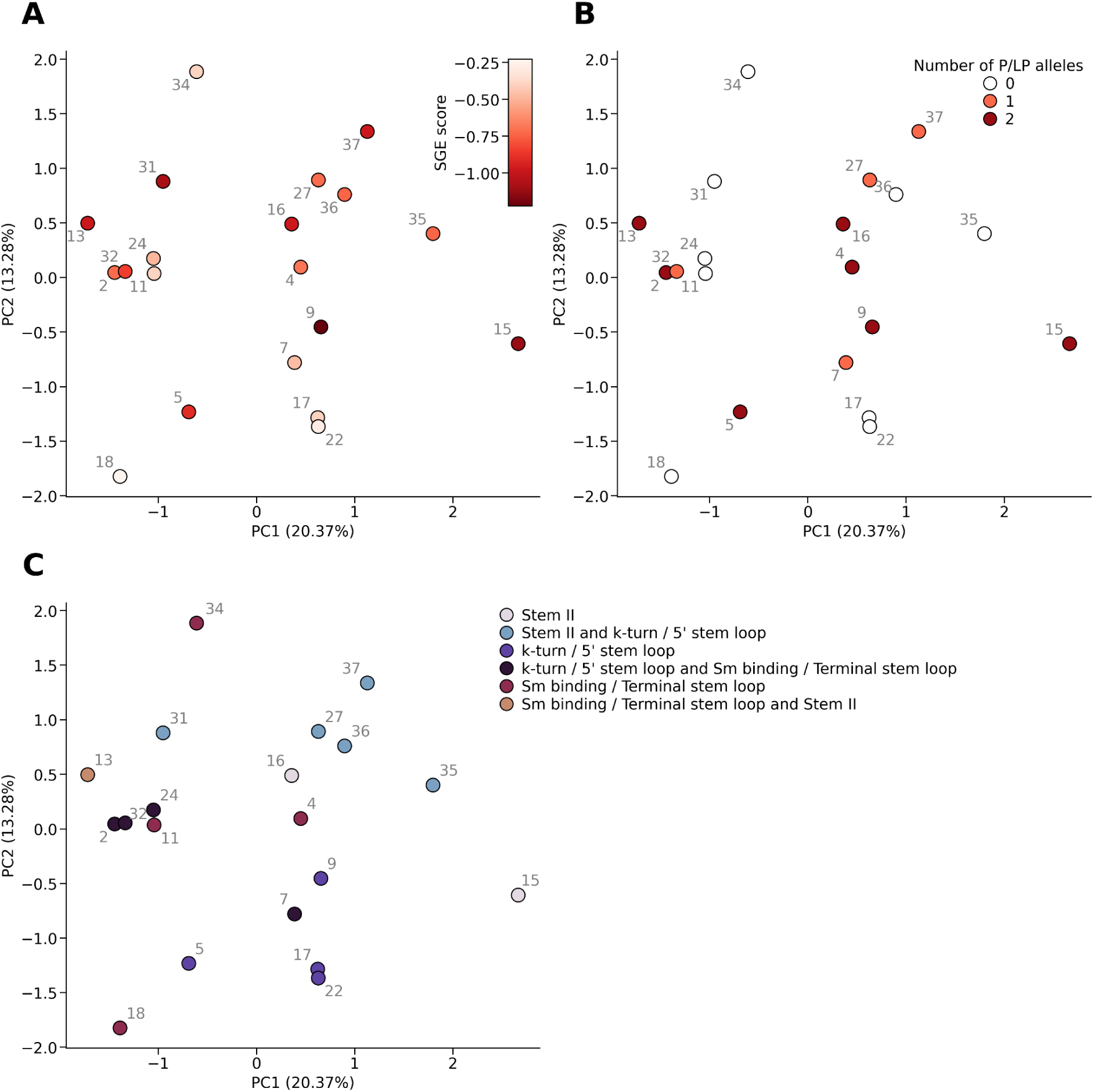
Principal components analysis for HPO terms in biallelic *RNU4-2* cases. (**A**) Points are coloured by the SGE score of the most deleterious variant in each individual. (**B**) Points are colored by the number of alleles with a Pathogenic / Likely Pathogenic classification. (**C**) Points are coloured by the position of each biallelic variant pair within the U4 secondary structure. The participant number is shown next to each marker in grey text. Only one individual from each sibling pair is shown. The percentage of the variance explained by each principal component is shown in the axis labels.

